# Systems Leadership: how Chief Executives manage tension between organisation and system pressures

**DOI:** 10.1101/2021.09.26.21264128

**Authors:** Ben Gordon, Matthew Gwynfryn Thomas, Lisa Aufegger, Ara Darzi, Colin Bicknell

## Abstract

**Aim:** System leadership is the requirement for a leader of a single organisation to operate on behalf of a wider system, rather than their individual organisation. It is not clear to what extent the current policy landscape supports leaders in managing misalignment between the needs of their organisation and the wider system, as many national structures still emphasise a focus on individual organisations. This study aims to understand how Chief Executives implement system leadership in practice when faced with decisions that benefit the system to the detriment of their own trust.

**Methodology:** Semi-structured interviews were conducted with ten Chief Executives from a range of trust types to understand their perceptions and decision-making process in practice. Semantic thematic analysis was used to draw out themes in relation to how Chief Executives approach decisions which weigh up the system and organisation.

**Results:** Themes raised by interviewees included both advantages (such as support in managing demand) and disadvantages (such as increased bureaucracy) of system leadership and practical considerations in operationalisation (such as the importance of interpersonal relationships). Interviewees endorsed system leadership in principle, but did not feel that the organisational incentives as currently structured support the implementation of system leadership in practice. This was not seen as a major challenge or impediment to effective leadership.

**Conclusion:** As a specific policy area, a direct focus on systems leadership is not necessarily helpful. Chief Executives should be supported to make decisions in a complex environment, without a specific focus on healthcare systems as a unit of operation.

## INTRODUCTION

Systems leadership is a concept that has been gaining increasing attention in many fields, particularly in organisational leadership in the public sector. For leaders to display systems leadership, they must think and act beyond the needs of a specific organisation, department or other entity for which they are responsible and instead act on behalf of a larger system. Defined by the NHS Leadership Academy in 2017, systems leadership is “…leadership across organisational and geopolitical boundaries, beyond individual professional disciplines, within a range of organisational and stakeholder cultures, often without direct managerial control.” [1]

The National Health Service (NHS) in England has many overlapping systems, arranged by geography, service type and clinical specialty. Organisations within systems are required to interact with each other in order to meet the needs of the patients they serve. As healthcare becomes increasingly complex, both technologically and operationally, the traditional approach of a patient receiving sequential care from a single organisation is no longer relevant, as a patient’s journey will often take them into contact, directly or indirectly, with multiple organisations. [2]

Given this arrangement, individual leaders could act purely in the interests of their own organisation, with the assumption that the ‘internal market’ arrangement would lead to high quality care by driving the organisations to act in the best interest of patients. However, it has been shown that holistic care across the health system is better for patients, as well as being more cost effective. [3]Therefore, there is a need for leaders in the health system to think and act in a more integrated way. There has been a move in recent years to increase the legislative footing of integrated care. [4] It has been suggested that this move has been accelerated to support recovery from the COVID-19 pandemic. [5]

The paradigm of a specific leader presiding over the priorities of a single organisation do not fit the needs of the constantly changing arrangements in England. The NHS is moving to a more collaborative approach to leadership through initiatives such as Integrated Care Systems (ICS). [6] Through these approaches, policy-makers wish to encourage holistic, collaborative thinking from Chief Executives. Organisations which form part of an ICS are encouraged to operate on behalf of the population, making decisions that benefit the organisations across the system, to the potential detriment of their own. [7]

However, there are several challenges in the move towards integrated care within England, including that of leadership structure. [8] Organisations are still presided over by leaders who remain accountable for their own organisations’ success. This means that Chief Executives are being pulled in two directions, on one hand being asked to operate on behalf of the system, beyond the boundaries of their organisation, while also being tasked with ensuring the stability of the organisation for which they are accountable. [9]

Many Chief Executives have cited fears being “decapitated by regulators” if their individual organisation fails to perform effectively. [10] Some have stated that the approach of national bodies such as NHS England, NHS Improvement and the Care Quality Commission is making things more difficult for leaders to operate in this way. [11] A key challenge to implementation is the tension between the benefit of the system, and the regulatory framework that incentivises a leader to protect their organisation. [12]

Existing literature explores the approach to implementing at a policy level, or in highlighting the challenges faced by individuals. [13] However little research has been conducted on how individuals approach the challenge. This research provides an evidence base for how individuals manage this challenge in practice.

## METHOD

### Interviews

Trust Chief Executives were chosen as the research subject group, as opposed to other executives or policy-makers, because as individuals they are specifically accountable for their own organisation, and are also required to support system leadership through Sustainability and Transformation Partnerships (STPs) or ICSs. Any subject who met the eligibility criteria (current or former Chief Executive) was deemed eligible for the research, provided an interview could be arranged. The study was reviewed and ethical approval given by the Joint Research Compliance Office, Imperial College London, UK (Reference: 19IC5317).

Semi-structured interviews were used, with a set of pre-defined questions and prompts. An initial set of questions were developed, informed by existing literature on system leadership, and shared with an academic working in the health policy space, a social researcher working for a large UK charity and a former NHS chief executive. Following this review, the questions were broadened in focus to a more general format, reflecting the absence of existing research on the practical management of the challenge to specify the details of the questions and further questions were added on demographics and the role of national bodies. This field test also provided prompts for the discussion. The final list of questions and prompts is given in Table 1.

**Table 1:**
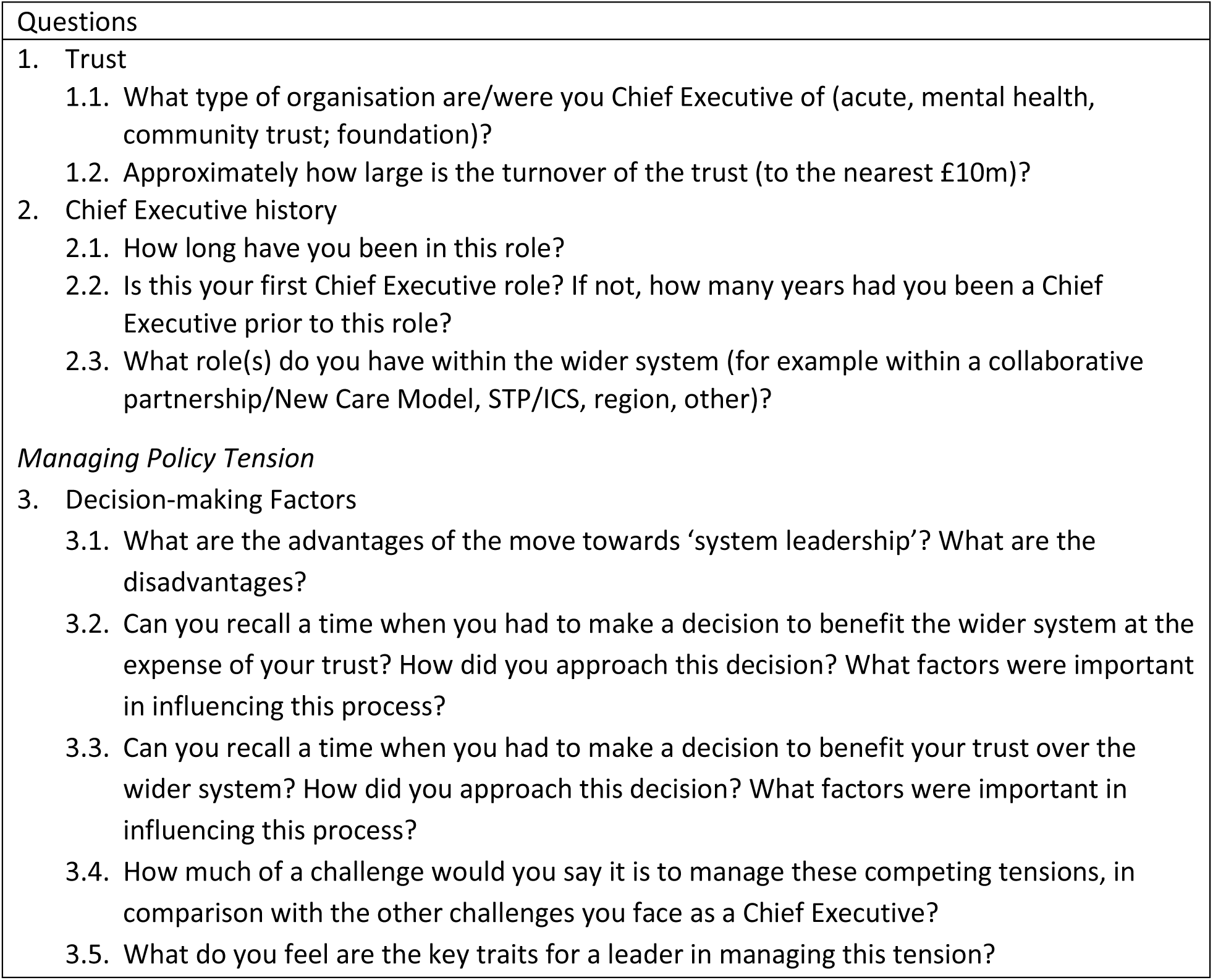
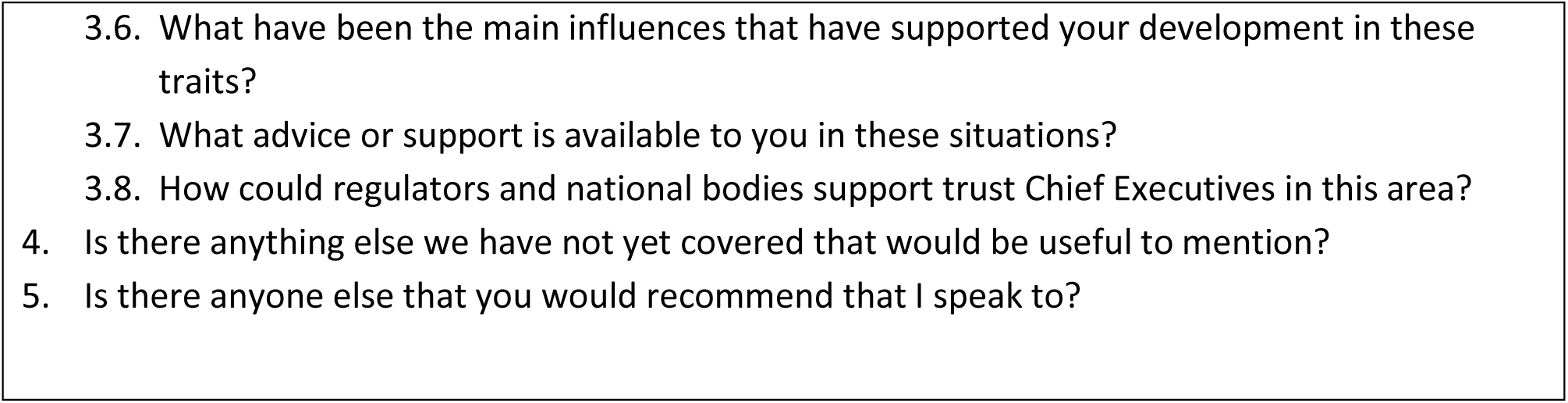
List of questions asked of interviewees

As a sampling strategy, a snowball approach was used to ensure wider coverage of interview subjects and increase potential diversity in interviewees. Emails were initially used to contact interviewees, in accordance with the ethical approval received for the project.

Interviews took place in the participants’ own time, and the interviews were a combination of phone calls and face-to-face, based on convenience and availability. Of the 10 interviews, 5 took place over the phone and 5 took place in person. Audio recordings were made to support analysis, following the interviewee’s consent. To improve the lines of questioning and subsequent analysis, ‘field notes’ were made during the discussion. Each interview lasted between 30 and 45 minutes.

### Data processing

The data collected was analysed using pragmatic thematic analysis. [14] The approach used for transcription was a form of content analysis, which begins the process of coding during transcription. [15] Following transcription, each transcript was read several times, and recurring concepts were highlighted and added to the initial emerging categories noted during transcription. The transcripts were then re-read with the emerging categories, and these were developed into identified themes. The themes were discussed with an independent researcher who undertook a separate analysis to confirm the thematic breakdown and review the strength of evidence for each. Digital analysis of word and word-pair frequencies was also used to review themes raised by researchers.

## RESULTS

There were ten Chief Executives interviewed, 3 from acute trusts, 6 from mental health trusts, 2 from community trusts and 1 from a specialist trust. The number of years’ experience averaged 6.6 years, ranging from 1 to 17. The average tenure of an NHS Chief Executive has been estimated as 2.5 years, and there is no available estimate of the average number of years’ experience in a Chief Executive role. [16]

### Themes

Analysis of the interviews raised thirteen sub-themes across four main themes, which are presented in Table 2 below, with each interviewee given a shorthand code and number (CEX)

**Table 2:** Themes and sub-themes raised by interviewees along with sample quotations

| Theme | Sub-theme | Sample Quotation | Number of interviewees raising topic |
| --- | --- | --- | --- |
| General | Discomfort with the term "systems leadership" | It's a term that's bandied around all over the place. Like the word integration it means different things to different people (CE2) | 4 |
|  | Balancing system and trust is not the most pressing issue to Chief Executives | it's a constant frustration, rather than an immediate worry. (CE9) | 8 |
| Advantages | Helps with demand through an economy of scale and co-operation | Now we look at how do we do things differently. Last week there was a discussion about shared corporate resources, like HR and IT, we're all spending on those things (CE7) | 4 |
|  | Helps target intervention to area of greatest need and root causes | the model of the NHS as just a national "sickness" service, which is what it was set up for, isn't really sustainable. So it has to be very good at treating the sick, that's still it's primary role, but it has to do more (CE5) | 7 |
|  | More support or buy-in to decisions made | If you do get a joint decision, everyone buys into this, and it's more likely to be sustainable and deliverable (CE3) | 1 |
| Disadvantages | The system does not support this way of working | the problem now is that we still judge people who are accountable for institutions, so we haven't really recalibrated the accountability and regulatory framework, but we are asking people to operate differently (CE3) | 7 |
|  | Increased administrative burden | You have to work really hard, lots of phone calls and meetings (CE6) | 5 |
|  | Indiscriminate implementation can harm organisations | If the amount of sharing of burden will only drag down the organisation that's performing well, that's still what we'll do (CE5) | 2 |
| Operationalisation | Importance of interpersonal relationships | It comes down to working with people (CE1) | 4 |
|  | Clear offers of support are needed from central agencies | What I'd like to see is [NHSE/I] being more collegiate and supportive, focusing on quality improvement rather than performance management (CE1) | 5 |
|  | Need to define success in advance | Without agreed outcomes, it becomes hard to make a change (CE2) | 4 |
|  | Role of board in decision-making | I haven't had any challenges from my board, saying you shouldn't be doing this and you should be pushing organisationally solvency. I think paradoxically NEDs arrive with a wider view than the organisation, as they're not in the organisation full time (CE8) | 6 |
|  | Consideration of local context | It depends on the position you're in. The bottom line is that every trust in the country is poor, no hospital is rich, it's just that some have significantly more problems than others (CE4) | 4 |
|  | Weighing up decisions that force choices between system and trust | (see Table 3 below) |  |

**Table 3:** Quotations relating to the “general” theme of system leadership

| Sub-theme | Quotation |
| --- | --- |
| Discomfort with the term “systems leadership” | It’s a term that’s bandied around all over the place. Like the word integration it means different things to different people. Everyone who says it implies that they’re a system leader. I’m very happy with the idea of working on a population focus. If you call that system leadership, and want to have outcomes, resource, finances and regulation lined up around that, that’s something I’m wholeheartedly in support of. (CE2) |
|  | One of the things I find, as a potential risk, is that people pepper all the dialogue with ‘system’ this and ‘system leadership’ and the system needs to sort this out, and it’s become almost a distraction (CE3) |
|  | We now wholeheartedly talk about system leadership in the same way that we jumped on the idea that we all have STPs and ICSs, and assume that everyone means the same thing, without asking about what it means for the person who’s accountable. (CE6) |
|  | I think the main concept, it’s a bit of a binary oversimplification to say we’ve had single organisation management at the expense of everything else. That’s partially true, especially if you look at something like the FT regime. But the reality is that all health and social care takes place in complex systems (CE8) |
| Balancing system and Trust is not the most pressing issue to Chief Executives | It’s helped me to work with the system – although I always have to keep one eye on my organisation because I don’t want to go to prison (CE1) |
|  | Not high up my list – given that we’ve had a long term plan in place and we’ve been asked to manage these challenges. (CE2) |
|  | It wasn’t top of the worry list (CE3) |
|  | that doesn’t particularly keep me awake at night. What disappoints me is that you can see what massive potential there is to do great things, what’s disappointing is the limitation that you have in doing this (CE4) |
|  | It’s the most often quoted thing by Chief Executives, in terms of anxiety. CEs who are responsible for institutions understand they are asked to be accountable for the community that they serve, but also the institution that they lead, and sometimes being a system leader means that you have to put some other player ahead of the best interests of your institution. (CE5) |
|  | Well it is a worry, I have to be honest. (CE7) |
|  | Actually this is not high up my worry list. It’s more like workforce – will I have enough staff for my services. That’s what worries me, as I suspect is the biggest worry for everyone. So this isn’t a worry, but it is a challenge to manage. (CE8) |
|  | ... it's a constant frustration, rather than an immediate worry. My immediate worry is: I want to make sure my services are safe, my A&E's don't collapse or that I've got no staff. But in terms of important things to change, it's right up there (CE9) |

**Table 4:**
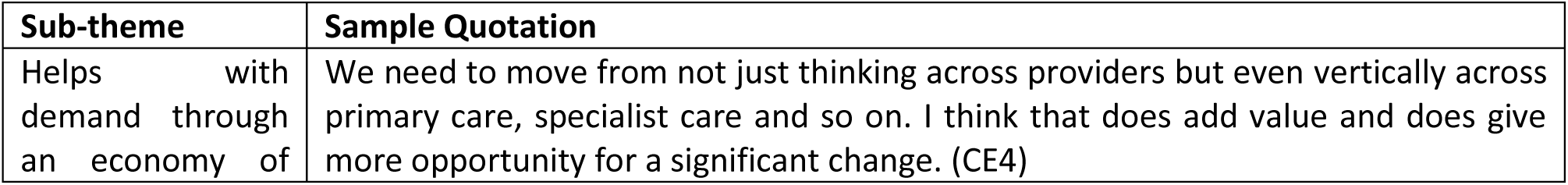
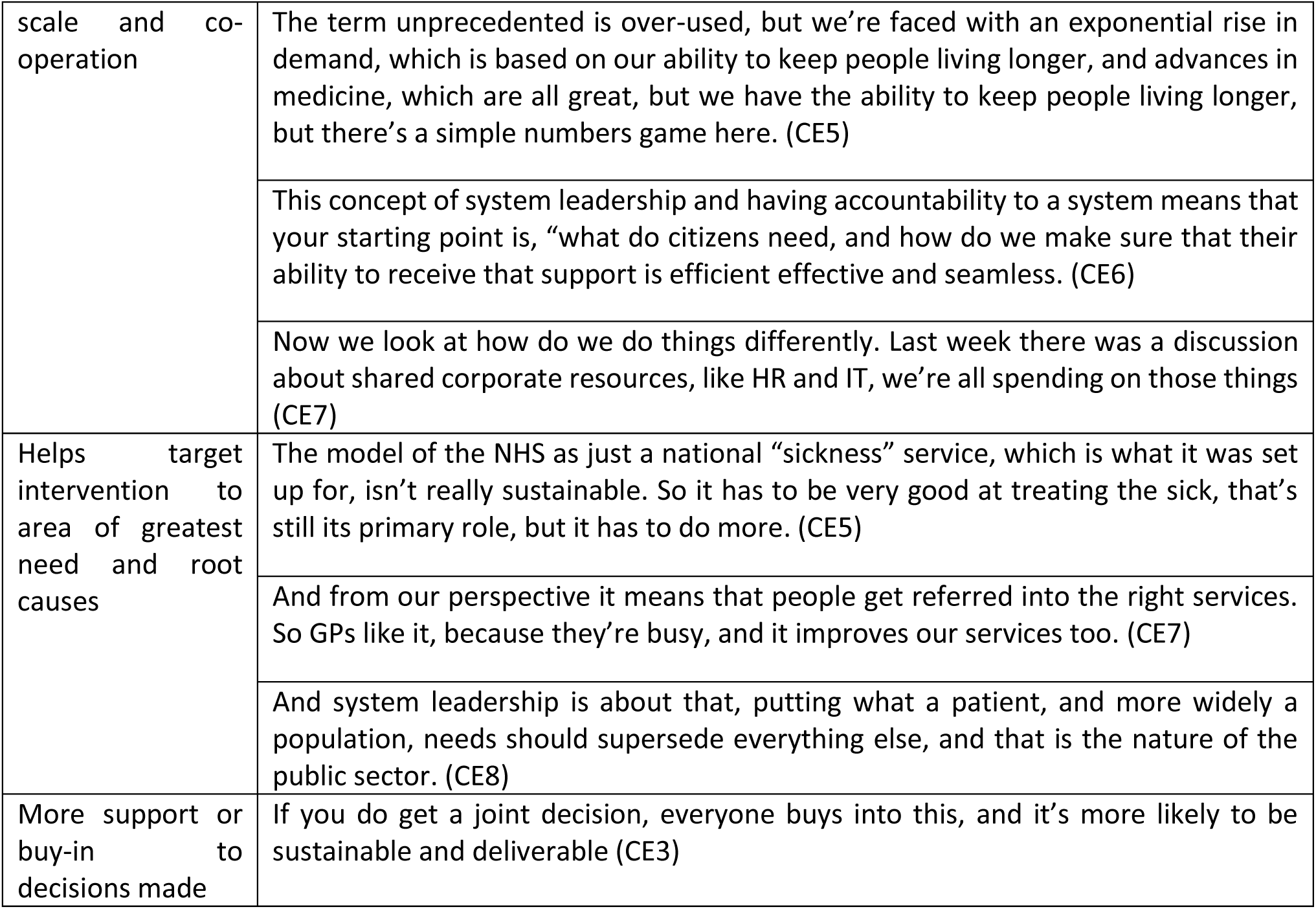
Quotations relating to the Advantages of System Leadership

### Sub-themes

#### General: Discomfort with the term “systems leadership”

While all participants favoured the underlying intention behind systems leadership, exemplified by one interviewee saying “no one can disagree with that.” For four interviewees, it was this very level of support that made it difficult for them to engage with the concept. Something as self-evident as the need for leaders to collaborate and operate on behalf of their local population did not warrant specific policies or attention. The lack of consistent definition and openness to reinterpretation was seen as a major challenge for implementation, because many felt that they had their own perspective of system leadership but that the idea held by others would be different.

#### General: Policy tension was not a major concern for most interviewees

When interviewees were asked how much of a challenge they felt the misalignment of incentives to be, only two stated that this was a major challenge. Most interviewees described it as a “frustration” that was not on their acute worry list – often citing workforce or safety concerns – but impacted their ability to operate strategically.

#### Advantage: Helps with demand through an economy of scale and co-operation

Four interviewees felt that system leadership would support them in achieving suitable economies of scale, which were seem as necessary given increasing demand on NHS services. Two used the term “common sense”, that it was obvious that this type of thinking was necessary if the NHS was to continue to meet resource constraints, related to the point above.

#### Advantage: Targeting interventions to greatest need and root causes

In addition to benefiting from an economy of scale, thinking about the health of the system, rather than an organisation or institution, supported people in thinking and operating in a preventative, public health-based way.

This perspective is supported by the digital analysis of word-pairs in the transcripts, which shows “mental health” and “social care” as two of the major topics raised by interviewees, suggesting that system leadership provides an opportunity to take a more holistic approach to healthcare (Figure 1).

**Figure 1:**
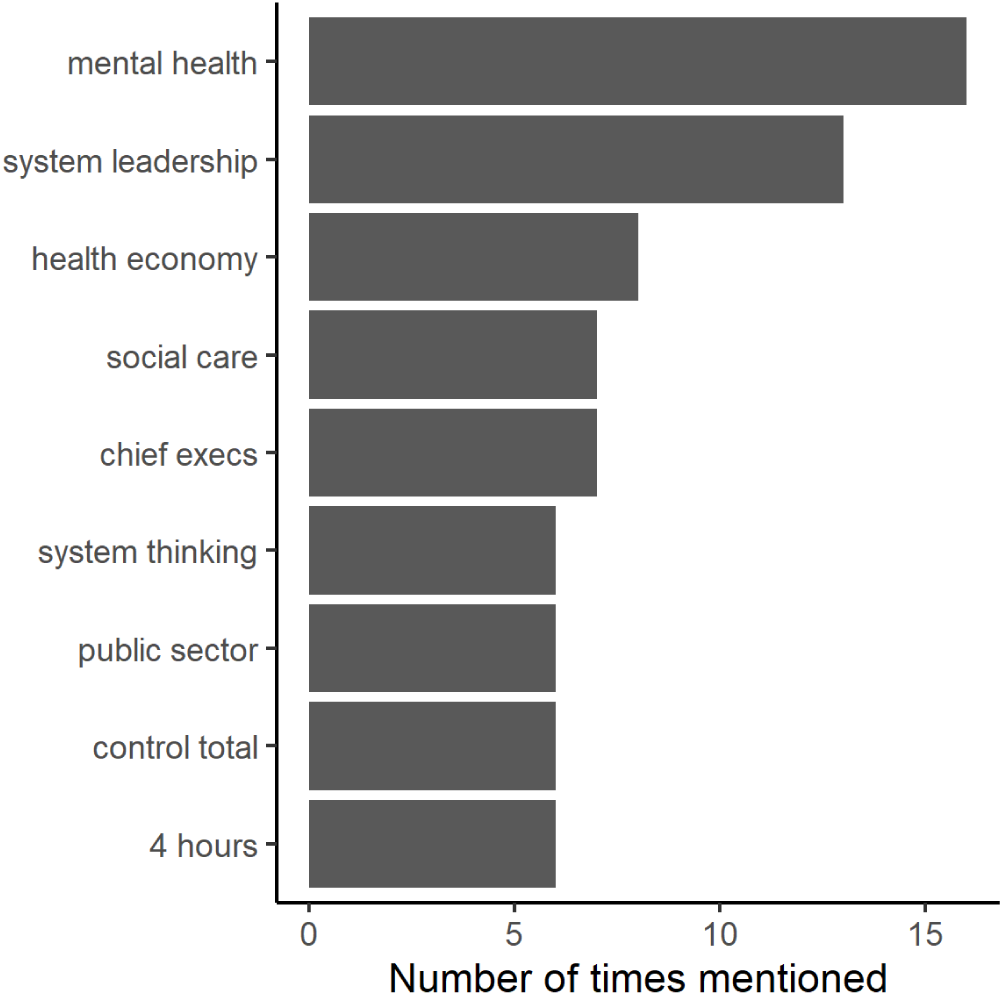
Frequency chart showing outputs of word-pair analysis of interview transcripts

#### Advantage: More support or buy-in to decisions made

One interviewee also noted that joint decision-making was likely to be more sustainable, and more parties were likely to buy-in to the decision that had been made.

There were also several disadvantages mentioned, however, the majority of these relate to the process of implementing systems leadership, rather than the concept itself. The responses relating to this theme are given in Table 5:

**Table 5:**
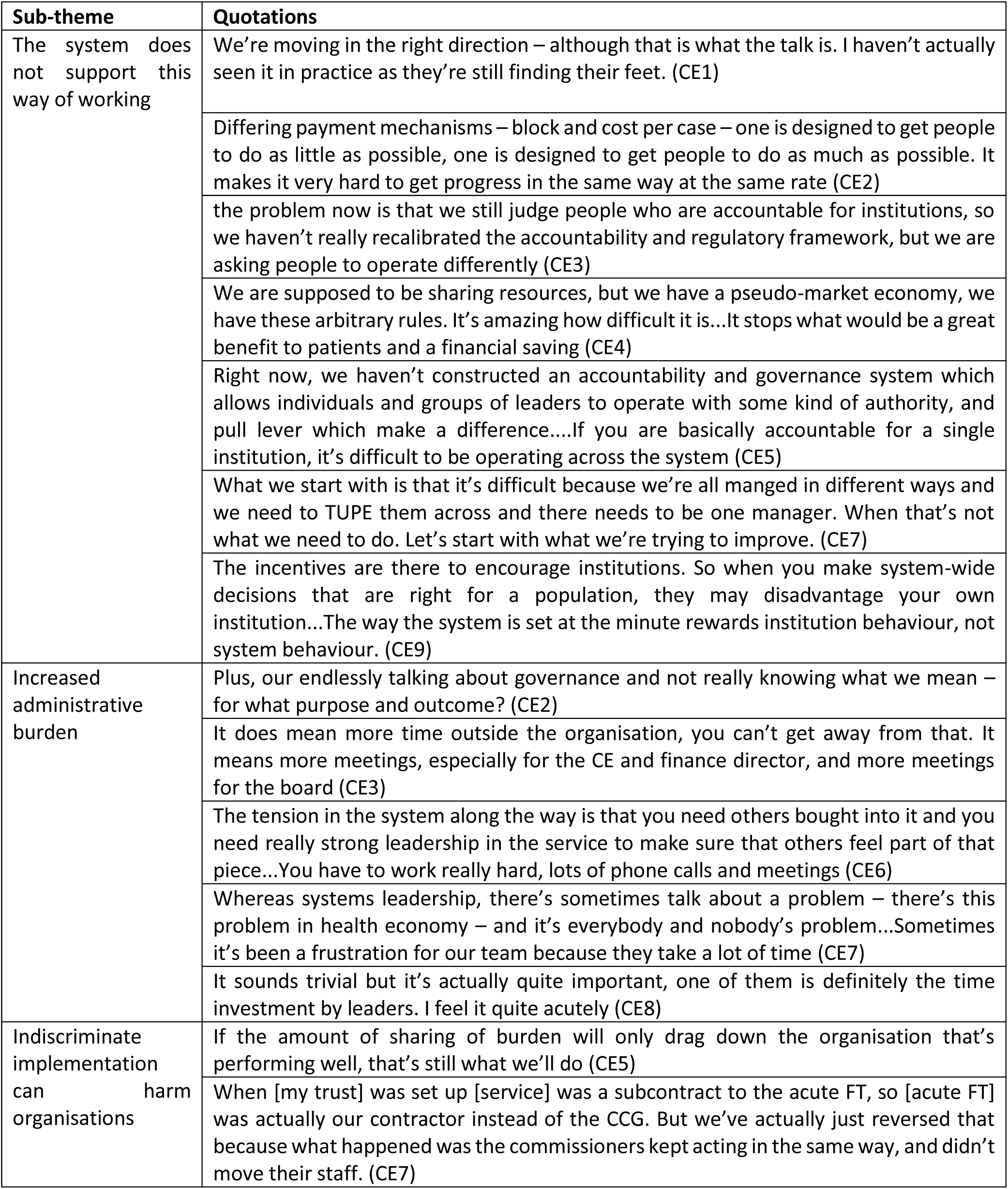
Quotations relating to the Disadvantages of System Leadership

#### Disadvantage: The system does not support this way of working

Many stressed how the overarching framework was not arranged in a way that supported systems leadership. This was seen in many areas, particularly finance, regulation, contracts and structures. Financial arrangements could interfere with systems leadership by blocking people from making specific decisions that would be beneficial for the system, or by incentivising different approaches due to differing arrangements between trusts. One interviewee brought up the challenges imposed by financial arrangements six times in the interview, at one point describing these as “fundamentally repugnant.”

#### Disadvantage: Increased administrative burden

Systems leadership was also seen as potentially more bureaucratic, placing a much greater burden on the time of senior leaders. While there is likely to be greater buy-in to the decisions made, as noted above, in order to achieve this, the process for making decisions will naturally take more time. Interviewees highlighted the amount of time spent on governance by NHS organisations.

#### Disadvantage: Indiscriminately implementing can harm successful organisations

According to interviewees, there are potential risks to successful organisations when implementing systems leadership. There is a danger that this approach can encourage ‘groupthink’ – if many trusts in an area are failing, operating more closely as a group could lead to the more successful ones lowering their standards.

Examples relating to the process of operationalising systems leadership are given in Table 6.

**Table 6:** Examples of decisions where an interview has had to choose between their trust and the system

| **Brief Description of example given by interviewee** | **Favoured...** |
| --- | --- |
| Decommissioned learning disability service at trust, to allow other trusts to provide the service | System |
| Took on lead role in a collaborative arrangement, including full share of potential gains | Trust |
| Adopted subcontractor role with another trust | System |
| Rejected subcontractor role after a period of time | Trust |
| Stepped back from providing a service to allow this to be taken on by a neighbouring trust | System |
| Accepted flat cash from commissioners to support increased funding to neighbouring trust | System |
| Pushed back on flat cash from commissioners for second year in a row | Trust |
| Purchased land as an STP asset | System |
| Negotiated contract settlement with commissioners due to increased demand | Trust |
| Agreed to publicly support commissioners' decision to fund an expensive service | System |
| Did not provide any contribution to system control total, due to financial pressures | Trust |
| Offered additional contribution to system control total to make up for other financially challenged trusts | System |
| Offered to provide additional scanning capacity to other trusts in the system | System |
| Agreed to join collective bank and agency pool across system, despite increased costs to trust | System |
| Making staff redundant to support a system objective | System |
| Consolidating a service with a neighbouring trust to improve quality | System |

#### Operationalisation: Importance of interpersonal relationships

Personal dynamics and interpersonal relationships were raised by all interviewees as being an important factor in implementing systems leadership. Many interviewees made reference to the fact that relationships with other parts of the system were very important, and that reciprocity was an important factor in decisions.

#### Clear offers of support are needed from central agencies

Many Chief Executives noted that NHS England and Improvement, as well as other agencies such as the Care Quality Commission (CQC) could adjust their approach to focus on supporting organisations, rather than punishing them for performance.

#### Need to define success in advance

A key element in implementation noted by interviewees was the process of identifying the definition of success in advance, and using this as a basis for decision-making. That way, a Chief Executive could be comfortable that even if, by certain metrics, their trust would not benefit, the decision would have a positive impact for clinical care in the wider health economy.

#### Role of boards in decision-making

Unsurprisingly, the majority of interviewees made reference to the role of the board in decision-making, particularly the Chair and Non-Executive Directors. These individuals have a key role in the implementation of systems leadership as they are responsible for the individual trust, and generally are less involved in discussions around the system, so can act as a ‘check’ for individuals undertaking systems leadership, or could provide a wider perspective supporting system thinking.

#### Consideration of local context

The status of the trust, in terms of its financial stability and quality rating, had a significant impact on decision-making. Three interviewees mentioned feeling comfortable about a decision that might benefit other parts of the system, at the expense of their own trust, because they felt confident that their trust remain solvent. Interviewees noted that “success” in the eyes of regulators and national bodies often led to more freedom from scrutiny and challenge, which supported the ability to implement systems leadership.

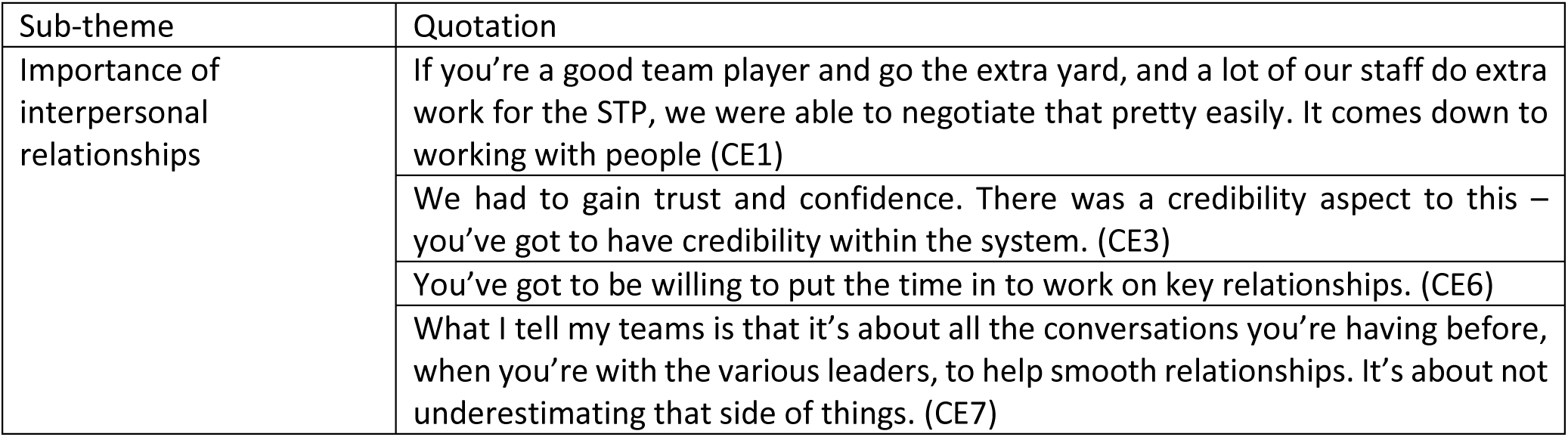
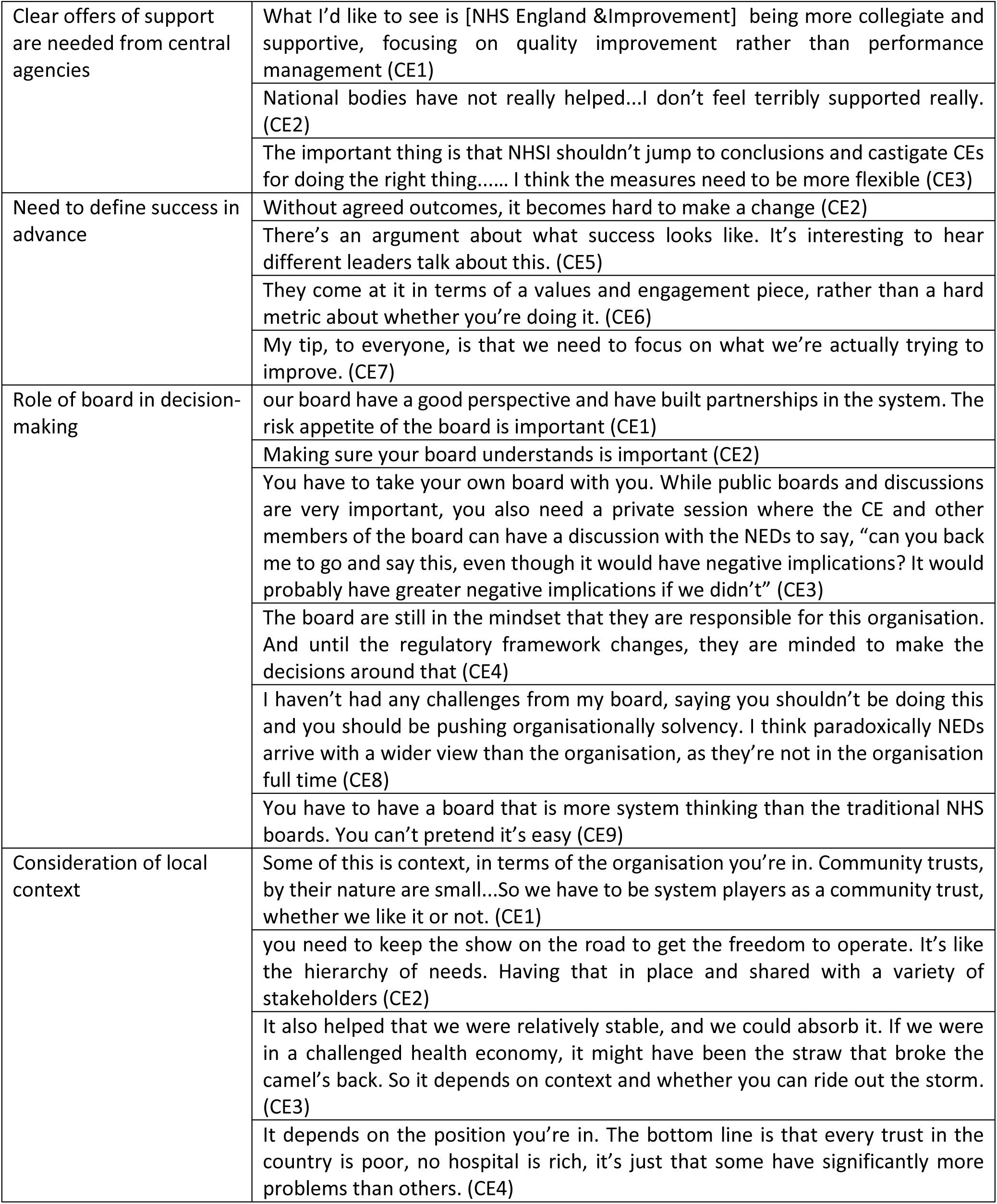

| Sub-theme | Quotation |
| --- | --- |
| Importance of interpersonal relationships | If you're a good team player and go the extra yard, and a lot of our staff do extra work for the STP, we were able to negotiate that pretty easily. It comes down to working with people (CE1) |
|  | We had to gain trust and confidence. There was a credibility aspect to this – you've got to have credibility within the system. (CE3) |
|  | You've got to be willing to put the time in to work on key relationships. (CE6) |
|  | What I tell my teams is that it's about all the conversations you're having before, when you're with the various leaders, to help smooth relationships. It's about not underestimating that side of things. (CE7) |
| Clear offers of support are needed from central agencies | What I'd like to see is [NHS England & Improvement] being more collegiate and supportive, focusing on quality improvement rather than performance management (CE1) |
|  | National bodies have not really helped...I don't feel terribly supported really. (CE2) |
|  | The important thing is that NHSI shouldn't jump to conclusions and castigate CEs for doing the right thing..... I think the measures need to be more flexible (CE3) |
| Need to define success in advance | Without agreed outcomes, it becomes hard to make a change (CE2) |
|  | There's an argument about what success looks like. It's interesting to hear different leaders talk about this. (CE5) |
|  | They come at it in terms of a values and engagement piece, rather than a hard metric about whether you're doing it. (CE6) |
|  | My tip, to everyone, is that we need to focus on what we're actually trying to improve. (CE7) |
| Role of board in decision-making | our board have a good perspective and have built partnerships in the system. The risk appetite of the board is important (CE1) |
|  | Making sure your board understands is important (CE2) |
|  | You have to take your own board with you. While public boards and discussions are very important, you also need a private session where the CE and other members of the board can have a discussion with the NEDs to say, "can you back me to go and say this, even though it would have negative implications? It would probably have greater negative implications if we didn't" (CE3) |
|  | The board are still in the mindset that they are responsible for this organisation. And until the regulatory framework changes, they are minded to make the decisions around that (CE4) |
|  | I haven't had any challenges from my board, saying you shouldn't be doing this and you should be pushing organisationally solvency. I think paradoxically NEDs arrive with a wider view than the organisation, as they're not in the organisation full time (CE8) |
|  | You have to have a board that is more system thinking than the traditional NHS boards. You can't pretend it's easy (CE9) |
| Consideration of local context | Some of this is context, in terms of the organisation you're in. Community trusts, by their nature are small...So we have to be system players as a community trust, whether we like it or not. (CE1) |
|  | you need to keep the show on the road to get the freedom to operate. It's like the hierarchy of needs. Having that in place and shared with a variety of stakeholders (CE2) |
|  | It also helped that we were relatively stable, and we could absorb it. If we were in a challenged health economy, it might have been the straw that broke the camel's back. So it depends on context and whether you can ride out the storm. (CE3) |
|  | It depends on the position you're in. The bottom line is that every trust in the country is poor, no hospital is rich, it's just that some have significantly more problems than others. (CE4) |

##### Managing the tension in practice

Interviewees were asked to discuss times they were faced with a decision that put the impact on the trust and the impact on the system at odds, and to provide examples of times they had favoured the system at the expense of their trust and vice-versa.

There were considerably more examples of decisions that benefit the system at the expense of the trust than the converse. When asked to provide examples of decisions that had encountered this tension, out of the 10 interviews, the interviewees mentioned a total of 11 examples where the Chief Executive had made a decision that benefited the system at the expense of the trust. There were only 5 examples of decisions to support the trust at the expense of the system. 5 interviewees were not able to provide an example of the latter.

The list of examples is given in Table 6:

## DISCUSSION

### Summary of findings

The interviews revealed a clear support for systems leadership amongst the interviewees, despite some uncertainty about the meaning of the concept, and whether others would interpret the meaning in the same way. The idea that leaders within the NHS should be mindful of broader needs than just their own trust and be focused on their own organisation was universally accepted, which is consistent with the existing literature about systems leadership in the NHS. [8]

While there was universal acceptance amongst interviewees that systems leadership is a valuable concept, there was also a clear consensus that the system does not support the application of system leadership, which is consistent with existing literature. [11] The disadvantages for systems leadership listed by interviewees were almost exclusively about the difficulty of implementing this concept in practice, and specifically the fact that the system itself did not support system leadership. This relates to the organisational frameworks and the financial and contracting mechanisms that limit and direct the responsibilities of Chief Executives in the NHS, and the regulatory framework that incentivises a focus on the success of the individual organisation. [7]

All interviewees noted that systems leadership is an important concept and that it is difficult to implement in the current system, but the majority of interviewees did not state this to be a major source of concern, instead citing challenges around workforce and waiting times as greater worries. Few interviewees cited the lack of support in implementing systems leadership as a major worry for them. It could therefore be concluded that, while implementing systems leadership in practice is a difficult task, it is no more challenging than many of the other difficult decisions facing NHS trust Chief Executives, such as managing workforce. While systems leadership may be a relatively new term, it does not introduce challenges that did not already exist. [11]

Almost all interviewees listed clinical benefit and the optimum outcomes for service users and the wider population as being a critical factor in decision-making. This appeared to be an ingrained approach to decision-making rather than something which had resulted from the increased emphasis on systems leadership. Whether in the capacity of an individual trust, or in relation to the wider system, Chief Executives are often faced with decisions where they aim to achieve the best outcomes for their population. There is a lot of overlap in the traits required in a leader of a system, as there are for a trust. The system can therefore support Chief Executives by reducing the level of complexity they face.

### Strengths and Limitations

This study may be the first example looking at the operationalisation of systems leadership through direct interaction with individuals required to implement this concept in practice. Detailed interviews with senior leaders in the NHS allow for a unique reflection on the act of implementing this in practice.

Given the status of current research, these findings are of value despite an unrepresentative selection of Chief Executives interviewed. Representation of trust types across the interview group does not mirror the distribution of trusts in the country. Acute trusts are underrepresented within the interview group, whereas mental health and community trust Chief Executives are over-represented.

One potential limitation is the fact that the interviews took place over mixed settings, with some happening over the phone and some face-to-face. This was a necessity given the limited availability of the interview subjects and time constraints of the research but may introduce bias relating to the level of detail or openness in a phone call compared to a face-to-face discussion.

The author’s role as an interviewer means that the author could unhelpfully prompt or direct interviewees. While this could introduce bias, it also has the potential to be beneficial to research, as it may draw out more information from interviewees. In grounded theory, the interface between data capture and theory means that having an interviewer and coder who is immersed in the theory allows the process to be fully interactive, and can lead to more informative results. [17] The role of the independent researcher in supporting the development of the themes has an important contribution to the rigour of the findings.

### Implications for health policy

Despite limitations listed above, the findings from this research are likely to be generalisable across other areas in the NHS. The consistency in views across interviewees, and the range of trust types and experiences suggest that this research is representative of views across many Chief Executives. The limitation relating to trust context (specifically, success and stability) may introduce some challenges, but other than this, it is likely that the findings could be more widely generalised. The implications for health policy are that:

- Systems leadership should not be viewed as a separate issue
- Policy-makers should aim to reduce complexity
- Central support should focus on supporting complex decision-making ahead of direct system leadership

Viewing “systems leadership” as merely another element of complexity, rather than a separate skill or way of working, means that the attentions of policy-makers should be focused on reducing complexity, and supporting Chief Executives to make difficult decisions. A direct focus on “systems leadership” as a standalone concept, paying attention to this area at the expense of others, can make implementation more difficult in practice. Creating bureaucratic approaches to supporting the system can increase the complexity and make operating in this environment more difficult, as noted by several interviewees who made reference to endless discussions about governance and a lack of clarity in decision-making.

Many interviewees alluded to the need for national bodies and regulators to be cognisant of the challenges in operating within the system, and support Chief Executives within this context, rather than taking a punitive view. Recognising the need for leaders to be trained in complex decision-making, rather than systems leadership, would support the relevant skills for success in this environment.

With increasing numbers of individuals being appointed formally to integrated care leadership roles and expected legislative support for integrated care, effective implementation of this policy will be vital if it is to have the desired effect.

### Further research required

This study only looked at the role of trust Chief Executives, and it would therefore be worthwhile to compare these views with other relevant leaders within the system, such as CCG Accountable Officers and leaders within councils and local authorities. It would also be beneficial to speak to those in a formal, appointed systems leadership role, specifically Integrated Care System Chairs, to compare these views with others less directly involved in the system.

As integrated care becomes more embedded within the NHS, it would be interesting to see how perceptions towards systems leadership change. As systems become a more standard environment for operating, will ‘systems leadership’ simply become ‘leadership’?

Finally, it would also be of value to explore how the approach to leadership within the NHS could be applied elsewhere. The interviewees in this study have shown a focus on overall benefits for the populations they serve, regardless of the impact on an individual organisation. It may be that this ‘utilitarian’ approach to leadership can provide insights elsewhere, and that other sectors could learn from how the NHS applies leadership.

## Conclusion

This research has shown that the concept of systems leadership, such as it is, is an unhelpful area of focus within the NHS. There are likely to be greater benefits from reviewing the structures of the system and supporting leaders within it to make the right decision by reducing the complexity they face. While the study covered a small sample of Chief Executives, the consistency of their views makes it clear that this conclusion can be applied more widely across the country.

## Data Availability

Thematic analysis available on request

## ACKNOWLEDGEMENTS & COMPETING INTERESTS

The authors gratefully acknowledge the contributions of all individuals who provided their input into this work through the interviews.

The authors declare no competing interests.

## CONTRIBUTORSHIP STATEMENT

BG planned the study, conducted the interviews and analysed the findings. MGT undertook a secondary, comparative analysis of the themes and the natural language processing of the results. LA supported the development of the findings and the drafting of the paper. CB and AD helped develop the questions and interview approach and overall methodology. The guarantor of the findings is CB.

